# Dysosmia and dysgeusia as differential diagnostics for clinical triaging of COVID-19 cases

**DOI:** 10.1101/2023.08.05.23293582

**Authors:** Pham Huu Thien Hoa Phong, Emanuele Brai, Aatmika Barve, Azarnoush Kouchiar, Jean-Marie Annoni, Lavinia Alberi Auber

## Abstract

Smell and taste disorders are recognized as frequent, and often the only, signs occurring in the early phase of SARS-Cov-2 infection and in many cases perdure as post-viral symptoms. This evidence raised a general reconsideration of chemosensory deficits, further suggesting that their appearance can be considered as a discriminative and predictive tool to detect COVID-19 cases. In this study, encompassing the first and second pandemic wave, participants estimated their olfactory and gustatory sensitivity, plus they were administered the validated Brief Smell Identification Test (BSIT). We observed that smell and taste impairments were mainly experienced by COVID-19-positive subjects with comparable severity of respiratory symptoms as non-COVID-19 patients. In addition, we noticed that the diagnostic power of subjective olfactory assessments upon SARS-Cov-2 infection is comparable to quantitative evaluation, suggesting that self-reporting could be adopted as the first line of intervention, anticipating more exhaustive procedures aimed at containing COVID-19 diffusion and consequently preserving general health. Overall, results from this work share similarity with other studies, therefore further underlying that olfactory and gustatory disbalance can be distinctive hallmarks in COVID-19 continuum.

## 1. Introduction

The severe acute respiratory syndrome coronavirus 2 (SARS-CoV-2) is responsible for the coronavirus disease 2019 (COVID-19), a pandemic identified at the end of 2019 in China that rapidly expanded worldwide [1]. COVID-19 symptoms range from life-threatening to moderate-mild signs. The most critical consequences are in the aged population with comorbidities, which further expose frail subjects to developing severe respiratory manifestations, eventually requiring hospitalization. It is now well-established that SARS-CoV-2 is not only an airways pathogens but also affects the central nervous system (CNS) [2–4]. The viral particles enter the CNS via the olfactory route [5–7] and can reach multiple brain areas triggering a heterogeneous spectrum of neurological symptoms. Among them, headache, tiredness, smell and taste alterations are experienced by more than 30% of infected subjects [8,9]. From the beginning of the pandemic, many publications reported COVID-19-induced olfactory and gustatory symptomatic [10–13], therefore reinforcing the hypothesis of a neurological comorbidity. Furthermore, smell and taste disorders coincide with up to 80% of COVID-19 cases [12,14–18]. All this evidence led the World Health Organization to list dysosmia and dysgeusia among the most common symptoms associated with SARS-CoV-2 (https://www.who.int/health-topics/coronavirus#tab=tab_3). Many works, from the first pandemic wave, found that smell and taste impairments appear during the early phase of the infection, being, in the instance of otherwise asymptomatic or paucisymptomatic cases, the only or recurrent signs observed [13,15,16,19–22]. Noteworthy, when COVID-19 started, the majority of reported chemosensory alterations were mainly based on subjective evaluations, largely consisting of written or phone surveys, without the support of objective tests or clinical diagnosis [12,14,15,19]. This lack was principally related to severe restriction measures, applied to contain the viral spread but that, on the other hand, heavily affecting also routine procedures, such as medical visits. However, despite the mere qualitative nature of smell and taste self-reporting, these sensory complaints contributed to better defining COVID-19 features, thus enhancing the understanding of its clinical course and the overall management of this global emergency.

Then, by mid-2020, coinciding with the timeline of this study (April-October), increasing studies collected data on smell and taste sensitivity both through auto-assessments and quantitative tests, therefore providing more complete and realistic outcomes about COVID-19-related chemosensory deficits [23–26]. Moreover, longitudinal studies about COVID-19-related dysosmia described that olfactory deficit can be reverted after several weeks from the SARS-Cov-2 infection onset. For instance, Moein et al., showed that 8 weeks after the first symptoms, 61% of COVID-19 patients (out of 100 cases) returned normosmic [27]. While since its appearance, SARS-Cov-2 variants have subdued the severity of COVID making the disease endemic in 2022, the chemosensory impairment remains a characteristic neurological trait along with the respiratory symptoms but also in the post-viral phase.

The main aim of this work was to assess whether and to what extent olfactory and gustatory impairments can be concomitantly experienced after SARS-Cov-2 infection and can be used alone to discriminate as the first line of diagnosis COVID cases among symptomatic subjects. We analyzed data on self-reported smell and taste disturbances from individuals showing mild to moderate respiratory symptoms that visited the COVID-19 emergency unit. In addition, the scores from subjective smell complaints were compared to quantitative evaluations obtained with the Brief Smell Identification Test (BSIT). Our observations indicate that dysosmia and dysgeusia are highly specific to COVID-19 in a population of patients with comparable respiratory disorders. This study is aligned with other works in describing olfactory and gustatory disturbances upon SARS-Cov-2 infection and further supports that these alterations, often experienced by paucisymptomatic or otherwise asymptomatic individuals, can be used as key differentials to identify COVID-19 cases and consequently contain the viral propagation to ensure public health.

## 2. Materials and Methods

### 2.1 Study participants

The recruitment was done between April and October 2020 and coordinated in specific COVID-19 areas within the Emergency Unit of the Cantonal Hospital of Riaz in the canton of Fribourg, Switzerland. 221 subjects arrived for acute respiratory complaints or conditions classified as from low, 0, to mild, 1, or moderate 2, with more than 90% having oxygenation support. All individuals were handed out the information leaflet and only interested participants signed the informed consent before their admission to this study. After, they underwent chemosensory probing and nasal swabbing for detection of COVID-19 positivity, via RT-PCR methodology (Cobas 5800, Roche). Among the 221 monitored persons, 39 resulted positive to SARS-Cov-2 and, from the remaining 182 COVID-19 negative, 79 were randomly selected, among the eligible group, to achieve a balanced comparison and avoid an over-representation of COVID-19-negative cases. The 118 participants were from both genders, with an age range of 17-90 years (Table 1). The exclusion criteria were: i) moderate to severe respiratory symptoms, ii) pregnancy and inability to give consent or follow procedures, and iii) insufficient knowledge of the project language. This research project was conducted in accordance with protocol CER-VD N.ID 2020-00695 respecting the Declaration of Helsinki, the principles of Good Clinical Practice, the Human Research Act (HRA), and the Human Research Ordinance (HRO) as well as other locally relevant regulations. The participants did not receive any monetary compensation for the study. 39 patients among the 221 were positive corresponding to the natural prevalence (21%) of COVID between the first and second waves in Switzerland in symptomatic patients (https://www.covid19.admin.ch/fr/overview).

**Table 1.** Summary of selected binary variables in the study cohort.

| VARIABLES | COVID_+ve | COVID_-ve | OR (95% CI) | z-score | p Value |
| --- | --- | --- | --- | --- | --- |
| N(F) | 79(39) | 39(21) |  |  |  |
| Age | 38.88±15.33 | 38.69±16.58 |  |  |  |
| Fever | 1.64±0.48 | 1.61±0.49 | 0.970 [0.542 , 1.734] | 0.104 | 0.458672 |
| Cough | 1.62±0.50 | 1.31±0.49 | 2.054 [1.148 , 3.674] | 2.426 | 0.007629 |
| Musc_pain | 1.75±0.43 | 1.92±0.26 | 1.554 [0.889 , 2.716] | 1.548 | 0.060794 |
| Diff-breathing | 1.51±0.50 | 1.35±0.48 | 0.901 [0.473 , 1.749] | 0.284 | 0.388387 |
| Nasal-discharge | 1.53±0.50 | 1.38±0.49 | 2.222 [1.254 , 3.937] | 2.734 | 0.003128 |
| Fluctuation_in_Smell | 1.46±0.68 | 1.49±0.73 | 0.709 [0.353 , 1.424] | 0.967 | 0.16678 |
| Quality of Smell | 2.03±0.96 | 2.00±0.97 | 2.982 [0.920 , 9.664] | 1.821 | 0.034297 |
| Sweet | 1.87±0.37 | 1.82±0.39 | 4.839 [2.389 , 9.805] | 4.377 | 0.000006 |
| Salty | 1.86±0.38 | 1.76±0.42 | 3.447 [1.741 , 6.824] | 3.551 | 0.000192 |
| Acid | 1.88±0.36 | 1.84±0.36 | 2.169 [1.011 , 4.652] | 1.988 | 0.023383 |
| Bitter | 1.88±0.35 | 1.82±0.38 | 2.169 [1.011 , 4.652] | 1.988 | 0.023383 |

These data, indicated as mean ± standard deviation, correspond to the variables reported in each cohort. (F)= females, OR=odds ratio, CI=confidence interval.

### 2.2 Questionnaire and smell test

Both cohorts filled out a French questionnaire (Supplementary data 1) indicating the presence (writing 1) or absence (writing 2) of multiple symptoms, such as fever, cough, muscular pain, nasal discharges, and also pathologies including diabetes, neurological, cardiovascular and respiratory disorders. Moreover, the examinees evaluated their olfactory and gustatory function using a scale from 0 to 10 points, with 0 indicating anosmia or ageusia and 10 normosmia or normogeusia. Additionally, the two groups underwent a validated smell test, namely the Brief Smell Identification Test [28] - a shorter version of the well-established University of Pennsylvania Smell Identification Test [29] - kindly provided by Prof. Richard Doty (director of the University of Pennsylvania’s Smell and Taste Center) and carried out with the assistance of a trained operator. This scratch and smell test, typically requiring 5 minutes, consists of 12 leaflets presenting a scent and 4 explicit odor identifiers with 1 correct choice. The final score was communicated to the participants and reported in the corresponding form (Baseline Clinical Data) for subsequent analysis.

### 2.3 Statistical analysis

Data collection and segmentation into two groups (COVID-19+ and COVID-19-) were executed in Excel 2022. We used R version 4.1.1 for all other analyses and graphics. We employed games_howell_test() to compare the group differences considering the violation of the assumption for homogeneity of variance. The results were considered statistically significant when the p-value was less than 0.05. Given the exploratory nature of the research, no first-order corrections were applied. Data were expressed in the text as average ± standard deviation. Graphical representation barplots were produced by ggplot() and likert plots were obtained by the likert() function of the ggplot2 package and HH and likert package respectively. We used corrplotfunc() function from the corrplot package to compare the spearman correlation in variables between COVID_+ve and COVID_-ve groups. For evaluating the diagnostic value of selected variables alone and in aggregate we performed sensitivity/specificity analysis respectively using MedCalc (https://www.medcalc.org/calc/diagnostic_test.php) and logistic regression modeling using the glm() function and analyzed the binary classifier using rocit() function and confirmed the sensitivity and specificity of the variables by calculating the area under the curve (AUC). These functions were utilized from glmnet and ROCit packages respectively.

## 3. Results

### 3.1 Cough and nasal discharge increase in COVID-19-positive subjects

Participants from both categories reported nasal discharge, muscular pain, cough, fever, and breathing difficulties as the most frequent symptoms (Figure 1A). Among them, cough (72% versus 53%, COVID_+ve = 1.30±0.52, COVID_-ve = 1.60±0.50, *p* = 0.004) and nasal discharge (69% versus 48%, COVID_+ve = 1.33±0.47, COVID_-ve = 1.53±0.50, *p* = 0.054) were strongly prevalent in the COVID-19 positive cohort compared to the negative one (Figure 1A). On the contrary, muscular pain (58% versus 46%, COVID_+ve = 1.43±0.49, COVID_-ve= 1.75±0.43, *p* = 0.33), fever (36% versus 35%; COVID_+ve = 1.64±0.47, COVID_-ve =1.64±0.48, *p* = 0.962) and breathing difficulty (23% and 24%; COVID_+ve = 1.76±0.42, COVID_-ve = 1.51±0.50, *p* = 0.908) showed a comparable prevalence between the two groups (Figure 1A). Next, we explored whether the enhanced nasal discharge experienced by COVID-19 subjects was associated with increased nasal obstruction. The collected responses pointed out that in both groups a similar percentage had nasal obstruction during the respiratory complaint (20.5% versus 22.8%, COVID_+ve = 7.61±2.68, COVID_-ve = 6.43+2.97, p = 0.841) (Figure 1B) and only few subjects in the COVID-19 negative group reported nasal obstruction before the onset of respiratory symptoms (0% versus 2.5%, COVID_+ve = 9.45±1.30, COVID_-ve = 8.94+1.84, p = 0.148) (Figure 1B).

**Figure 1.**
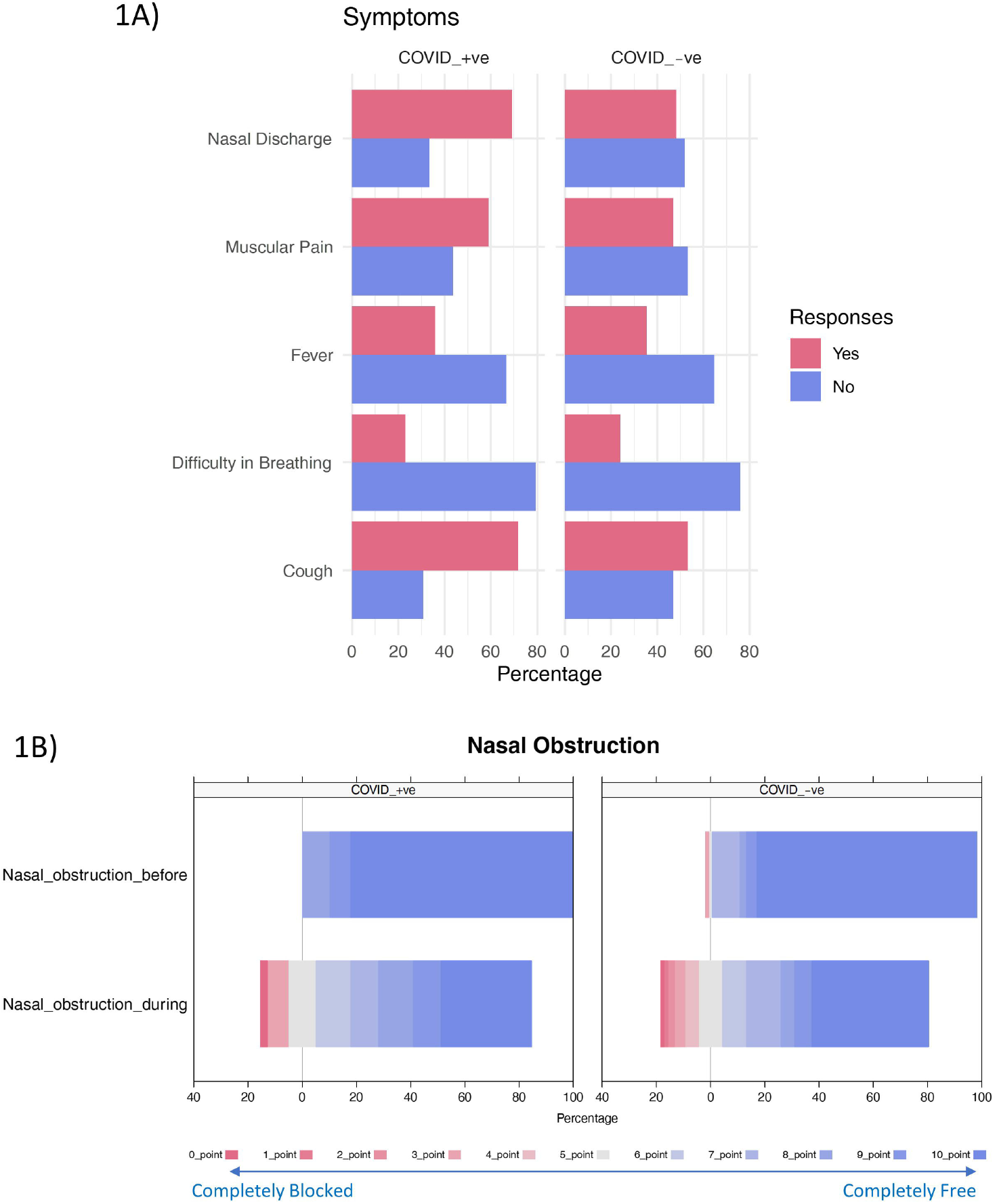
Changes in general symptoms and nasal obstruction in COVID-19 cohort. A) Differential representation of signs in COVID_+ve and COVID_-ve subjects includes: Nasal Discharge (COVID_+ve = 1.33±0.47, COVID_-ve = 1.53±0.50, *p* = 0.054); Muscular Pain (COVID_+ve = 1.43±0.49, COVID_-ve= 1.75±0.43, *p* = 0.33); Fever (COVID_+ve = 1.64±0.47, COVID_-ve =1.64±0.48, *p* = 0.962); Difficulty in breathing (COVID_+ve = 1.76±0.42, COVID_-ve = 1.51±0.50, *p* = 0.908); Cough (COVID_+ve = 1.62±0.46, COVID_-ve = 1.31±0.5, *p* = 0.007). B) Change in nasal obstruction before and during COVID-19 infection auto-evaluated with a 0-10 point scale (Completely blocked - Completely free) (COVID_+ve = 7.51±2.54, COVID_-ve = 7.61+2.68, *p* = 0.841).

### 3.2 Olfactory impairment is exacerbated in COVID-19-positive subjects

We addressed whether olfaction could vary in the two groups by self-reporting. First, we noticed that in both clusters more than 60% of examinees did not show any fluctuation in smell (Figure 2A), but those suffering from COVID-19 described a more pronounced alteration than controls (25.6% versus 19%), (Figure 2A). The remaining 10% of COVID-19-positive and 15% of negative individuals were uncertain about the answer (Figure 2A). Second, the quality of smell was significantly lower in COVID-19 patients in contrast to the negative group (82% versus 44%, COVID_+ve = 1.30±0.64, COVID_-ve = 2.03±0.96, p = 0.0000084) (Figure 2B). Third, auto-evaluation of smell perception during the symptomatic period revealed a diminished smell perception in COVID-19 cases compared to the negative ones (33% versus 17%; COVID_+ve = 6.66±3.9, COVID_-ve = 7.98±2.3, p = 0.05) (Figure 2C), whereas before manifestations there was no difference between groups (Figure 2C). Fourth, we determined olfactory ability administering the BSIT test, which underlined more severe hyposmia in the SARS-Cov-2 infected strata, where almost 36% of individuals, against 20% of the controls, totalized low score ranges (0-2 points) (COVID_+ve = 2.92±1.45, COVID_-ve = 3.48±1.23, p = 0.04) (Figure 2D).

**Figure 2.**
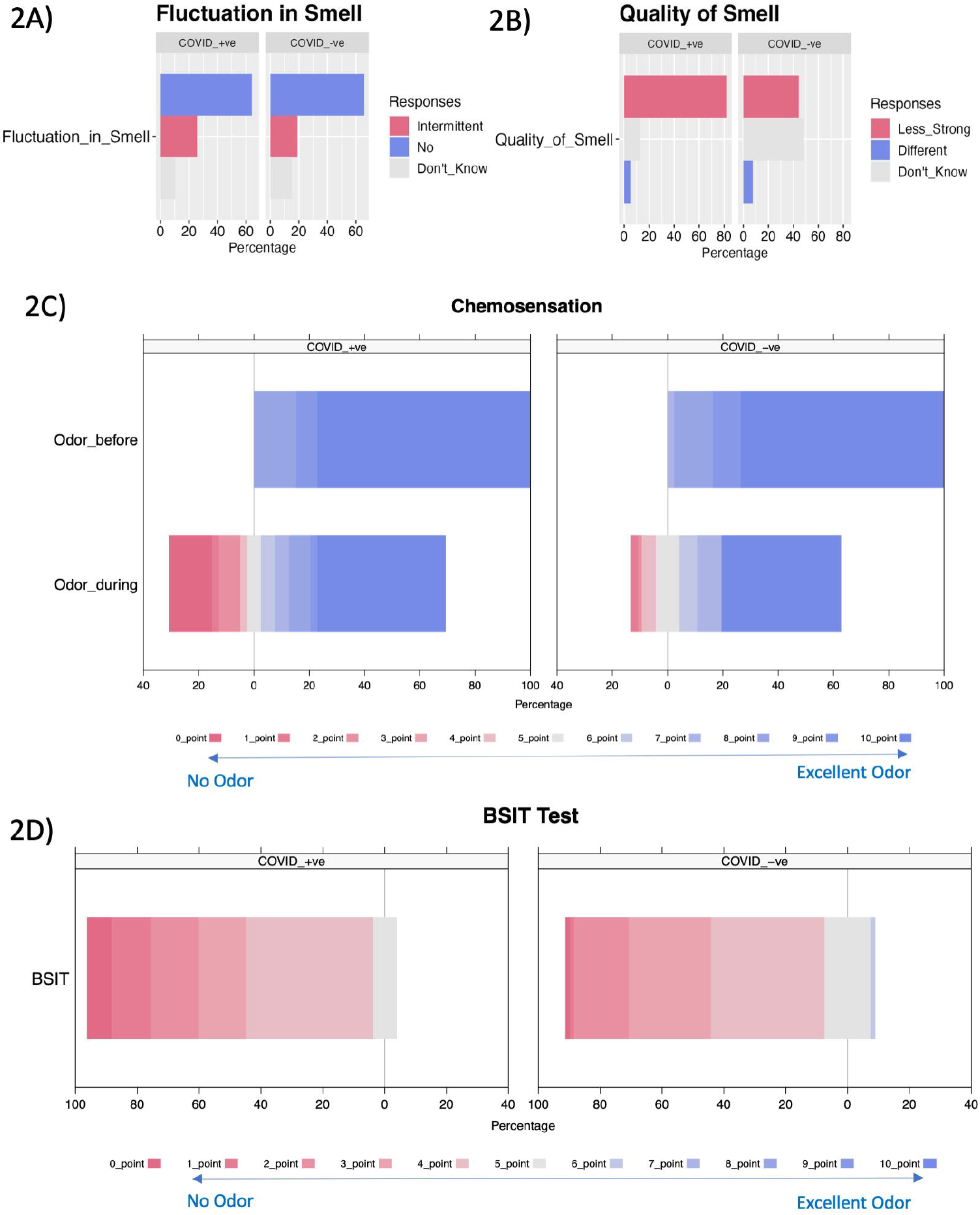
Exacerbated olfactory sensitivity in COVID-19 cases. A) Difference in smell fluctuation between COVID_+ve and COVID_-ve patients that auto-evaluated this aspect as: “intermittent”, “no change”, or “don’t know” (COVID_+ve = 1.46±0.68, COVID_-ve = 1.49±0.74, *p* = 0.816); B) Difference in smell quality between COVID_+ve and COVID_-ve patients that auto-evaluated this aspect as: “less strong”, “different”, “don’t know” (COVID_+ve = 1.30±0.64, COVID_-ve = 2.03±0.96, *p* = 0.000008); C) Change in chemosensation during COVID-19 infection self-evaluated on a 0-10 point scale (No odor - Excellent Odor) (COVID_+ve = 6.66±3.9, COVID_-ve = 7.98±2.3, *p* = 0.05); D) Subjective olfactory variation was validated using BSIT assessement (COVID_+ve = 2.92±1.45, COVID_-ve = 3.48±1.23, p = 0.04).

### 3.3 Taste alterations prevail in COVID-19-positive subjects

Beyond olfactory disorders, also dysgeusia was more experienced in COVID-19 individuals compared to the control counterpart (Figure 3A). We observed that positive subjects displayed a poorer taste sensitivity (between 0 and 5 points) than negative ones (30% versus 13%; COVID_+ve = 6.51±3.74, COVID_-ve = 8.55±2.18, *p* = 0.003) (Figure 3A), while no difference was reported before the onset of respiratory disturbances (Figure 3A). Moreover, participants specified whether and which taste type - sweet, salty, bitter and acid - was mainly affected. COVID-19 patients had significant taste changes in sweet (41% versus 14%; COVID_+ve = 1.58±0.49, COVID_-ve = 1.87±0.37, *p* = 0.03) and salty (38% versus 15%; COVID_+ve = 1.61±0.49, COVID_-ve = 1.86±0.38, *p* = 0.008) compared to negative individuals (Figure 3B). On the contrary, bitter and sour alterations, despite more frequent in the COVID-19 positive group, (bitter, 23% versus 13%; COVID_+ve = 1.76±0.42, COVID_-ve = 1.88±0.35, *p* = 0.146) (sour, 23% versus 13%; COVID_+ve = 1.76±0.42, COVID_-ve = 1.88±0.35, p = 0.146) didn’t show major differences between the two populations (Figure 3B).

**Figure 3.**
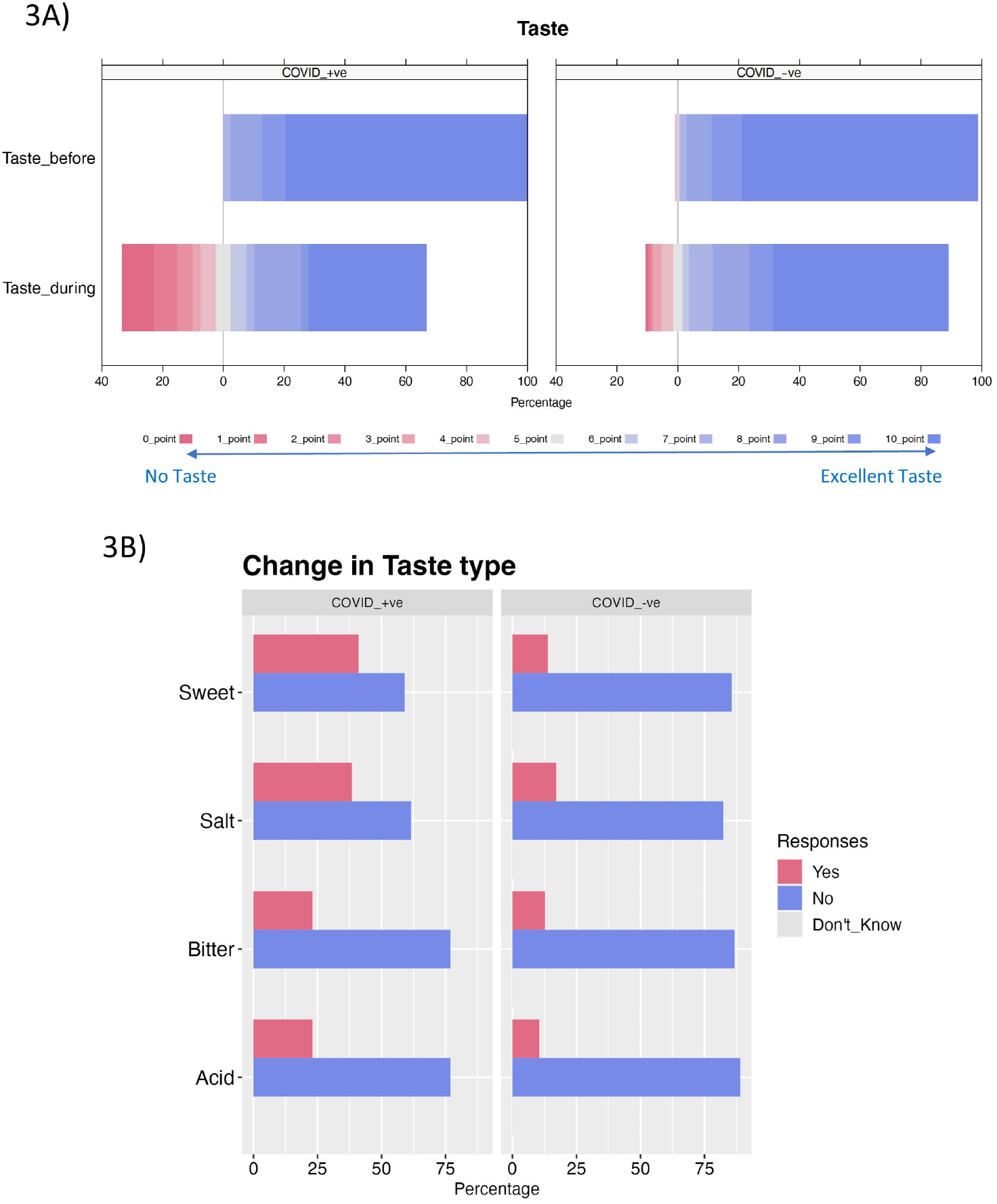
Taste alteration during COVID-19. A) Change in gustatory perception during COVID-19 auto-evaluated on a 0-10 point scale (No Taste - Excellent Taste) (COVID_+ve = 6.51±3.74, COVID_-ve = 8.55±2.18, *p* = 0.003); B) Change in taste types auto-evaluated using: “yes”, “no”, “don’t know”. Sweet (COVID_+ve = 1.58±0.49, COVID_-ve = 1.87±0.37, *p* = 0.03); salty (COVID_+ve = 1.61±0.49, COVID_-ve = 1.86±0.38, *p* = 0.008); bitter (COVID_+ve = 1.76±0.42, COVID_-ve = 1.88±0.35, *p* = 0.146); acid (COVID_+ve = 1.76±0.42, COVID_-ve = 1.88±0.35, p = 0.146).

### 3.4 Co-occurrence of smell and taste alteration in COVID-19-positive subjects

We performed a correlation analysis to better understand whether and which primary symptoms appear simultaneously during SARS-Cov-2 infection and then explored if those variables can be considered as diagnostic co-morbidities. Remarkably, the strongest association observed in COVID-19 subjects was related to self-reported hyposmia and dysgeusia (R=0.9, p<0.01), with a lower magnitude in the control group (R=0.67, p<0.01) (Figure 4A). Also, the correlation between olfactory dysfunction during the infection and changes in all taste types was higher in positive individuals (R=0.59 - 0.73) compared to the negative ones (R=0.4 - 0.44) (Figure 4A). To establish the diagnostic value of the continuous testing variables, namely i) self-reported smell, ii) taste rating and iii) BSIT, we run a sensitivity and specificity analysis. All variables demonstrated low sensitivity in identifying true positive COVID-19 cases (33-36%), but higher specificity for detecting COVID-19-positive patients versus controls (80-86%) (Table 2). We also interrogated whether using those three tests together would allow us to rapidly isolate COVID-19 cases. Receiver Operating Curve analysis indicated that performing the 3 aforementioned tests increases accuracy (AUC empirical = 0.695, AUC binormal = 0.702, AUC nonparametric = 0.677) (Figure 4B). Interestingly, when only considering the self-reported chemosensory deficit the accuracy is comparable (AUC empirical = 0.684, AUC binormal = 0.699, AUC nonparametric = 0.674) (Figure 4C), supporting the use of auto-evaluations. To further assess the diagnostic value of aggregating the 3 versus 2 variables, we conducted the Kolmogorov-Smirnov test (KS) which shows a higher differentiating capacity of utilizing the three continuous variables as compared to two in distinguishing COVID-19 positive from COVID-19 negative cases (Figure 4D and 4E).

**Table 2.**
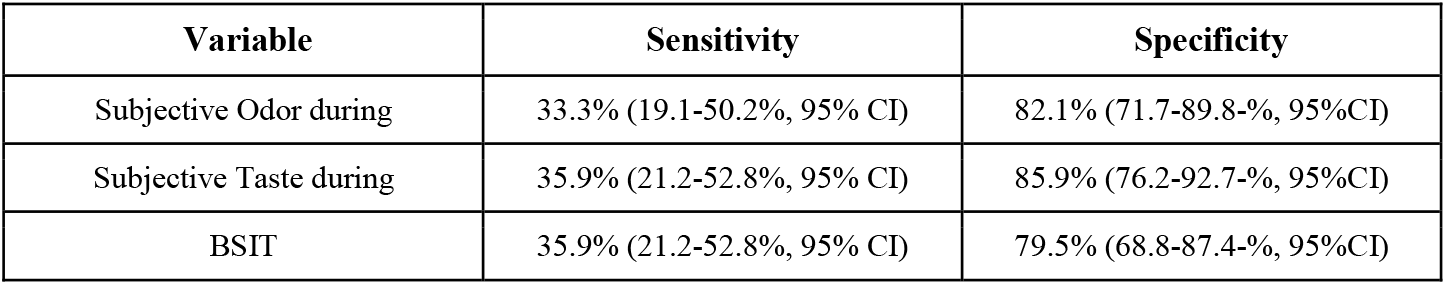
Summary of Specificity and Sensitivity analysis of selected variables with a scale.

**Figure 4.**
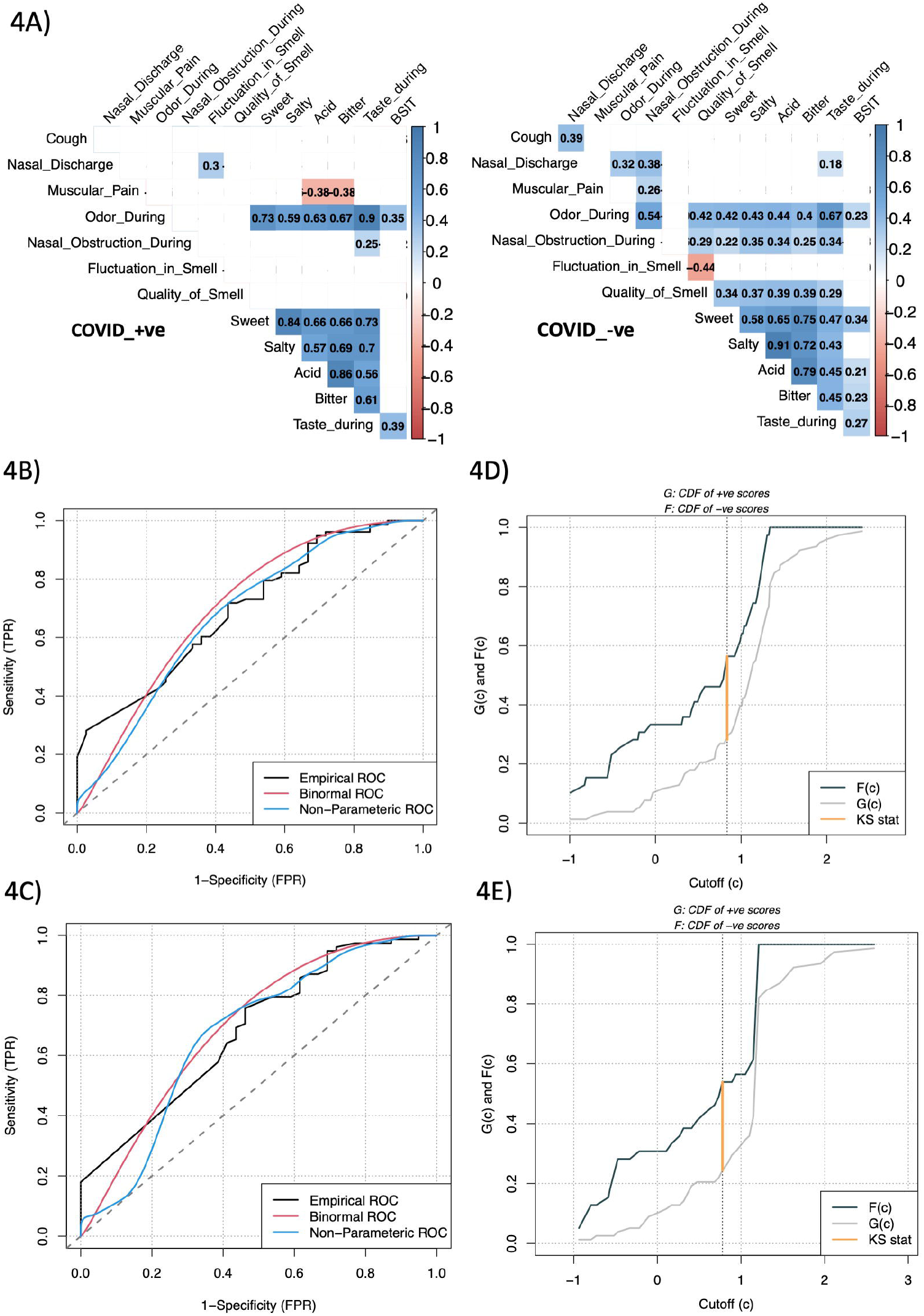
Dysosmia and dysgeusia strongly correlate in COVID-19 cases, showing a better diagnostic power. A) Correlation values for the variables between COVID_+ve and COVID_-ve cohorts. Maximal correlation in “Odor during” and “Taste during” (COVID_+ve, R = 0.9, p <0.01) in COVID-19 affected group as compared to the control one (COVID_-ve, R = 0.67, p <0.01). Stronger correlation between “Odor_during “ and “BSIT” in COVID_+ve group (R = 0.35, p < 0.01) and also between “Odor_during “ and “Quality_of_smell” (R = 0.63, p < 0.01). Only significant values (p < 0.05) are represented in the correlation plot. Receiver Operating Analysis for diagnostic value of B) Odor_during, Taste_during and BSIT and C) Odor_during and Taste_during. The plot represents three kinds of ROC curves on the binary classifier for the variables: Empirical, Binormal and Non-Parametric. The dotted line in the curve represents the “Random chance line”. Here, ROC curves represent the ratio between True positive rate and the False positive rate for the selected variables to diagnose COVID-19 positivity. KS plot for diagnostic value of D) Odor_during, Taste_during and BSIT and E) Odor_during and Taste_during. The plot represents the power of differentiation of the aggregated variables.

## 4. Discussion

The global and rapid escalation of SARS-COV-2 infection in 2019 caused a long-lasting socio-economic crisis that affected our society in multiple sectors. During the first pandemic wave, when this study started, the healthcare system of many countries collapsed due to the high number of hospitalizations triggered by respiratory insufficiency and its sequelae. In particular, subjects with pre-existing pathologies were the most vulnerable categories, due to the systemic effects of SARS-Cov-2 on the immune, and cardiovascular systems but also the brain. As of today, thanks to improved strategies in managing this emergency, the vaccination campaign, and the naturally occurring less virulent forms of the virus, such as Omicron, COVID-19 is no longer regarded as a severe condition but an endemic respiratory disease. The currently prevalent Omicron variant was first isolated in South Africa in November 2021 (https://www.who.int/activities/tracking-SARS-CoV-2-variants). Compared to the previous lineages, Omicron displays higher transmissibility and shares similar but milder reactions, such as fever, cough, nasal congestion, and sore throat, but preserves olfactory and gustatory deficits, making those symptoms very specific to SARS-Cov-2 infections independently of the variant.

Indeed, smell and taste alterations triggered by COVID-19 were frequently observed from the first wave, where either multicentric or single studies described these symptoms [13–15,19]. Moreover, these dysfunctions often occur in the early phase of the disease [13,30] and could be the only detectable manifestations [19,31], therefore making them a valuable target for the identification and isolation of paucisymptomatic subjects and so important for containing the viral diffusion. Consequently to this pandemic, smell and taste disruptions, already widely associated with neurological conditions such as dementia or depression [32–34] are gaining progressive attention, since they could potentially anticipate and be related to CNS sequelae captured in the long-COVID carriers [25,35–38].

In this study, conducted between the first and the second pandemic wave in 2020, we aimed to extract differential self-reported and objectively-assessed symptoms able to discriminate between COVID-19 positive and negative subjects with the scope of facilitating triaging and isolation of potential cases before testing. Among the self-evaluated signs, cough and nasal discharge were stronger in COVID-19-positive individuals, whereas fever, muscular pain, difficulties in breathing and nasal obstruction didn’t differ in the two groups. Nevertheless, in isolation, the majority of COVID-19-positive subjects reported cough (72%), nasal discharge (69%) and muscular pain (59%), which is in good accordance with previous reports worldwide [15,39,40]. Interestingly, nasal discharge due to mucus hypersecretion is associated with the acute cytokine storm caused by SARS-Cov-2 infection [41], which might affect also nasal flow and result on the associated olfactory and gustatory disturbances without a prevalent nasal obstruction. Overall, the adverse reactions we observed were also described in other works [13,42], indicating that the symptoms triggered by this virus are similar in subjects monitored in different centers and countries and also at different time points along the epidemiological evolution of the disease supporting the specificity of these respiratory symptoms.

In addition to the specific respiratory condition, we evaluated whether smell and taste could be used as differentials among a population of subjects with mild to moderate respiratory symptoms. Subjective and objective smell assessment both indicated that SARS-COV-2 affected olfaction in a very specific way, with self-reported smell loss concomitant to a drop in quality of smell (82%, R=0.63, versus 44% in seronegative patients). Hyposmia was then confirmed using the psychophysical test BSIT, indicating a marked deficit in COVID-19 patients compared to negative subjects (35% versus 20%). Furthermore, our study shows that subjective and objective smell deficits measured using BSIT are in good accordance (33% versus 35%). This is in contrast to other studies that report a higher degree of smell deficit using quantitative measures [43] and suggests that results are highly dependent on the type of test used as well as the ethnicity and geography.

We noticed that, along with the olfactory deficit, and consistent with other works [13,44], personal estimation of taste capacity pointed out that SARS-COV-2 strongly exacerbates this sense in COVID-19 cases against negative individuals (35% versus 13%). Furthermore, and in line with other studies [26,45,46], we observed that SARS-CoV-2 affected taste types. In particular, they were impaired with the following order of magnitude: sweet, salty, bitter, and sour. Sweet showed the strongest differentiating value (OR=4.9) within the binary variables followed by salty (OR=3.4). Thus subjects self-reporting concomitant hyposmia with cough (OR=2.1), nasal discharge (OR=2.2), drop in the quality of smell (OR-2.9) and dysgeusia are more likely to be at risk for COVID-19 infection.

While this data is historic, the chemosensory symptoms accompanying COVID-19 also in its milder form could be explained by the shunting of olfactory neuroepithelium and taste buds. These deficits are in most cases reversible except for long-COVID haulers, where they may represent an early asymptomatic sentinel for later neurological sequelae [37].

Limitations of this work are the relatively small sample size, in particular for the COVID-19 positive group, as well as the low statistical power, thus the obtained results, despite being in line with other evidence, should be cautiously interpreted and would require further confirmation. Moreover, other reports have raised the issue of unexplained intra- and inter-subject variability, a factor whose influence on the current data cannot be ruled out. Another feature to be acknowledged is that this study focuses on a particular cohort of patients, recruited among those arriving at the emergency unit because of respiratory complaints. Also, targeting different olfactory tasks, such as odor threshold and discrimination, as well as the administration of validated taste assessments, would render this evidence more robust.

In conclusion, we noted that, in accordance with other epidemiological studies, olfactory and gustatory impairments occur upon SARS-COV-2 infection. Furthermore, the diagnostic value of auto-reporting smell and taste alterations in SARS-Cov-2 was comparable to objective smell identification tests supporting the use of self-reporting as the first line of detection [15] making them a valuable target to monitor and control the viral diffusion, in concomitance with other strategies, to eventually preserve personal and public health.

## Data Availability

All data produced in the present study are available upon reasonable request to the authors

## Author contribution statement

Pham Huu Thien Hoa Phong, conducted clinical trial and patient diagnosis/survey/testing; Emanuele Brai wrote and revised the manuscript, checked the raw data and contributed to the conceptualization of the figures; Aatmika Barve analyzed the data, prepared the figures and contributed to the writing of the results; Azarnoush Kouchiar, supervised the clinical trial; Jean-Marie Annoni and Lavinia Alberi designed the clinical trial and reviewed the manuscript. All authors provided the final approval for the manuscript submission.

## Acknowledgment

This study has been started with the mission of adding evidence-based knowledge to the Global Consortium of Chemosensory Research (GCCR), an open science collaboration initiated to coordinate worldwide crowdsourced research aimed at understanding the reports of the chemosensory issues. The authors are grateful to Prof. Richard Doty, director of the Penn Smell and Taste Center at the University of Pennsylvania, for his insightful comments and advice during the manuscript preparation.

## Funding

The study was supported by intramural funding from the Cantonal Hospital of Fribourg and the Swiss Integrative Center for Human Health.

## Declaration of competing interest

The authors declare that they have no known competing financial interests or personal relationships that could have appeared to influence the work reported in this paper.

## Notes

### Competing Interest Statement

The authors have declared no competing interest.

### Funding Statement

This study was funded by intramural fund of the Cantonal Hospital of FR and the EKSAS fellowship from the Swiss Confederation.

### Author Declarations

On April 27th 2020, the Ethics Cantonal Commission of Vaud (CERVD) gave ethical approval for this work under the ID license: 2020-00695

