## Supplementary data for "Dysosmia and dysgeusia as differential diagnostics for clinical triaging of COVID-19 cases"

COVID-19 **Relationship between loss of smell & loss of taste**

Please complete this short questionnaire to find out how COVID-19 and non-COVID respiratory symptoms affect your sense of smell and taste. Please fill in as much as you can.

N° Patient Code:	Hospital: RIAZ, Fribourg

Your information :

| Today's date : |
| --- |

| Age : |  | Gender : | Men |  | Woman |  | Other |  | Don't want to tell |
| --- | --- | --- | --- | --- | --- | --- | --- | --- | --- |

| How were you diagnosed with COVID-19? | Symptoms |  | Viral test |  | Don't know |
| --- | --- | --- | --- | --- | --- |
| Date of symptoms : |  |  |  |  |  |

| Do you feel cured? | Yes - Completely |  | Yes -Partially |  | No |  | Don't know |
| --- | --- | --- | --- | --- | --- | --- | --- |

| What symptoms do you have? | | |
| --- | --- | --- |
|  | | *Duration (days) :* |
| *Fever* |  |  |
| *Cough* |  |  |
| *Breathing difficulties* |  |  |
| *Nasal discharge* |  |  |
| *Muscle pain* |  |  |
| *Other* |  |  |
| *If you answered Yes to Other, specify the symptoms :* | | |
| Do you have any of these diseases? | |  |
| *Diabetes* |  |  |
| *Hypertension* |  |  |
| *Heart disease (e.g. former heart attack)* |  |  |
| *Respiratory diseases (e.g. asthma/chronic emphysema)* |  |  |
| *Neurological diseases (e.g. stroke)* |  |  |
| *Chronic rhinosinusitis (e.g. sinusitis/polyps)* |  |  |
| *Other* |  |  |
| *If you answered Yes to Other, specify the symptoms :* | |  |

Sense of smell :

| Can you estimate your : | | |
| --- | --- | --- |
|  | Sense of smell : | Breathing through the nose: |
|  | *Draw a line* | *Draw a line* |
| *Before illness :* |   0 10  *No sense of smell Excellent* |   0 10  *Completely blocked Completely free* |
| *During illness :* |   0 10  *No sense of smell Excellent* |   0 10  *Blocked completely Completely free* |
| *After recovery: (if possible)* |   0 10  *No sense of smell Excellent* |   0 10  *Blocked completely Completely free* |
| Since the disease, the sense of smell is : | |  |
| *The sense of smell is weaker than before* |  |  |
| *Smell is different (e.g. quality of smell has changed)* |  |  |
| *Don't know* |  |  |

| Since the disease, your sense of smell has fluctuated? |
| --- |
| *No - no change* |
| *Yes - it comes and goes* |
| *Don't know* |

Taste :

| Since your illness, have you noticed a change in your ability to smell the : | | |
| --- | --- | --- |
|  | | *Duration (days) :* |
| *Sweet* |  |  |
| *Salted* |  |  |
| *Sour/acidic* |  |  |
| *Amer* |  |  |

| Can you estimate your : | |
| --- | --- |
|  | Sense of taste :  (for sweet, salty, sour and bitter only) |
|  | *Draw a line* |
| *Before illness :* |   0 10  *No taste Excellent* |
| *During illness :* |   0 10  *No taste Excellent* |
| *After recovery: (if possible)* |   0 10  *No taste Excellent* |
